# Hematological Parameters as Predictors of COVID-19 Infection: A Cohort Study Using Brazilian Hospital Data

**DOI:** 10.1101/2024.05.08.24307015

**Authors:** James Utley

## Abstract

**Objectives:** To determine if hematology test results can indicate COVID-19 infection.

**Methods:** A cohort study used 2020 Brazilian hospital data for 6,573 patients to run inferential analyses of fourteen red and white blood cell hematology test results, adjusted for age and sex, by patient COVID-19 status.

**Results:** Ten hematological biomarkers were statistically significant predictors of COVID-19 PCR test results; the strongest were Total White Blood Cells: OR=0.90 (0.88-0.93), Basophils: OR=0.97 (0.97-0.98), Total Red Blood Cells: OR=0.88 (0.78-0.99), Hemoglobin: OR=0.93 (0.89-0.97), Hematocrit: OR=0.97 (0.96-0.99). Patient sex was also a strong predictor of COVID-19 PCR test results: OR=1.43 (1.26-1.62).

**Conclusions:** COVID-19 produces measurable impacts on red and white blood cells that are significant predictors of PCR results. A routine blood draw may offer probabilistic insight as a diagnostic predictor of infection. The virus is associated with impacts in red blood cells’ vital oxygen transport attributes.

**Public Health Implications:** A faster diagnostic tool for COVID-19 will assist providers to triage and isolate those with the virus, reducing hospital transmission. Results are being used to develop a predictive model with high sensitivity and specificity.

## Introduction

The SARS-CoV2 virus is identified using polymerase chain reaction methods (PCR) to confirm COVID-19 infection. Healthcare systems rely on PCR testing to diagnose patients first presenting with symptoms to determine proper triage and treatment. However, PCR testing requires extensive resources that many developing regions lack.^1^ Even the wealthiest countries experience supply shortages due to continued high demand and production limitations. A molecular PCR lab requires stable electricity, supplies, reagents, sterile workspace, a sequencer, and trained technicians to run the analyses and record the results accurately.^2,3^ PCR testing also requires time for samples to be analyzed and results returned to the healthcare provider for action. The COVID-19 test takes 24 hours or more to fully process with PCR, causing an automatic day delay for diagnosing patients.^4^ Delays in diagnosis can cause inaccurate risk stratification during triage of first-present patients with suspected COVID-19 symptoms.^5,6,7,8^

Countries with the greatest population densities tend to have less resources available for their public health systems.^3^ Brazil in particular was severely impacted early in the pandemic during the time of this study. The 2020 W.H.O. figures report 1,368,195 confirmed cases, and 58,314 deaths from COVID-19 in Brazil; highest levels in the world only after the U.S.^10^ Brazil has limited PCR testing capabilities to satisfy the demand, which limits healthcare workers ability to accurately identify and isolate COVID-19 patients.^11,12^

When patients first-present at a healthcare facility reporting COVID-19 type symptoms, they meet the criteria for a PCR test to be ordered, if available. The percent of incoming patients testing positive accounts for a relatively small percentage of the total patient load; reported rates for COVID-19 positive results based on PCR testing range from 8% to 25% in 2020.^13^ This statistic alone underscores the importance of a fast indicator of COVID-19 infection because most ill patients being admitted to hospitals do not have the virus and should be safely segregated as soon as possible from the smaller percentage of those with the virus.

Patients being seen at nearly all healthcare facilities have their blood drawn for routine laboratory diagnostics and a Complete Blood Count (CBC) is conducted to produce quantified measures of the hematology profile, which include biomarkers for Red Blood Cells (RBC) and White Blood Cells (WBC). Quantified results are compared to a validated reference range to determine if abnormal levels are present in the patients’ blood. The processing time for CBC results is faster than PCR testing; a STAT specimen can be processed and available to the ordering provider within 60 minutes.^14^

Although COVID-19 is contracted through the respiratory system, viral replication and immune modulation manifests in the tissues and blood through SARS-CoV-2 virions entering host cells via mediated-endocytosis.^16^ The process is similar to other viral infections such as predicate Coronavirus’, Influenza, and Respiratory Syncytial Virus.^17^ The SARS-CoV-2 has a unique rapidly mutating spike protein that is responsible for interacting with host cellular membrane receptors including ACE2, CD147 and NRP1. All three receptors are now suspected to be involved in virus infection and replication.^16,18^

Changes in blood levels for specific WBCs (Leukocytes) is a typical immunological response to pathogens because the cells are deployed to combat infection. However, COVID-19 appears to stimulate some novel differential immunological responses in WBC biomarkers, as well as unexpected changes in certain Red Blood Cell (RBC) biomarkers, when compared to a typical viral infection.^19,20^

Leukocytes are nucleated cells that are a central component of the body’s protective immune mechanism.^20^ Different studies have reported abnormal responses in certain Leukocytic biomarkers in their COVID-19 patients.^21^ Lymphopenia was the most common finding throughout the literature.^7,22,27^ Research suggests lymphocytes are infected and injured, decreasing the circulating lymphocytes.^7^ Basophil and Eosinophil counts have also demonstrated significance in the COVID-19 disease continuum both in acute and recovery phases.^23,24^ Eosinophilia has also been reported in patients with COVID-19.^25,26^ Neutrophilia was reported in two studies.^2,12^ Monocytes have not appeared frequently in the literature in regards to COVID-19, aside from the ability for SARS-CoV-2 infected monocytes having the ability to produce inflammatory factors that aid in the COVID-19 cytokine storm, yet it is uncertain that the absolute count is affected in any way.^21^ Thrombolytic events and Platelet dysfunction were reported in advanced COVID-19 patients; collectively, studies report thrombocytopenia as a finding.^27,28^ Most of these studies were conducted with a small sample of late-phase COVID-19 positive patients.

Changes in Red Blood Cell counts (Erythrocytes) is not typical during viral infections because they are not active participants in adaptive or innate immune response systems. Erythrocytes are the most abundant cells in blood; they are highly flexible (as they must bend and twist as they pass through the capillaries) biconcave disks with the primary and critical function to transport oxygen, and to a lesser degree, carbon dioxide in the blood.^29,30^ Only a few studies have reported abnormal findings in some RBC biomarkers in COVID-19 patients.^29,31,32^ Severe anemia has been reported in some COVID-19 patients.^1^

Age of patients has been a clear contributor to COVID-19 infection rates since the start of the pandemic.^33^ The elderly community has been hit hardest; children had much lower susceptibility to the early strains.^34^ Along with age comes comorbidities, which have been shown to be associated with severe illness and death for COVID-19 patients.^12,35^ The effect of a patient’s sex is unclear regarding the virulence of COVID-19, but there are descriptive reports that males are testing positive at a higher rate compared to females.^1,9^

### Purpose

The purpose of this study was to analyze if results within the fourteen biomarkers measured in a routine Complete Blood Count can be a predictive diagnostic tool of COVID-19 infection. The hypothesis is that the virus leaves a unique biosignature in the blood that can be detected in the CBC results of patients, even in the early phase of infection.

## Methods

This epidemiology study used a cohort design with retrospective secondary data. The de-identified data was made publicly available through the COVID-19 DataSharing/BR initiative for the Hospital Israelita Albert Einstein in Sao Paulo, Brazil.^10^ Data are from November 2019 to October 2020, prior the Delta and Omicron variants. The database contains CBC test results for patients with and without COVID-19, as determined by PCR testing. The Joint Commission hospital is accredited, and practices COVID-19 PCR testing quality standards as required by the United States and the W.H.O. Inclusion criteria required subjects have data for their Complete Blood Count and their definitive COVID-19 PCR result from the time when they first-present with symptoms. For subjects with multiple sets of results, only the first set of CBC results were used for these analyses. Selecting data only for first-present patients allowed for the examination of biomarkers in the early phase of the disease.

The dependent outcome for the study was the PCR test result for COVID-19, reported nominally as either positive or negative. The independent predictive variables examined were the fourteen biomarkers measured in a routine CBC order. The CBC test measures each component of the hematology profile, including seven biomarkers related to erythrocytes, and seven biomarkers for the leukocytic cell lines, all reported at the interval level of measure. The erythrocytic components include Total Red Blood Cells, Hematocrit, Hemoglobin, Red Cell Distribution Width, Mean Corpuscular Volume, Mean Corpuscular Hemoglobin Concentration, and Mean Corpuscular Hemoglobin. The leukocytic biomarkers consist of Total White Blood Cells, Lymphocytes, Basophil Count, Neutrophil Count, Monocyte Count, Platelet Count, and Eosinophil Count. Two demographic variables were considered; patient age was measured at the interval level, and patient sex was measured nominally.

Multivariate logistic regression analyses were conducted to determine the potential predictive value of using CBC results as an indicator of COVID-19 infection. Inferential analyses were run on each of the fourteen hematology biomarkers, adjusted for age and sex, to determine their predictive value on the outcome of the PCR test result.

## Results

Raw data from electronic health records for 44,879 patients were downloaded from the data sharing website. Files were reduced to the subjects who met the defined study criteria. The study sample consisted of 6,573 subjects with data for all study variables at the time they first presented at the hospital with symptoms. The final set of study data were validated by the source document. Assumption tests were conducted for each study variable for normality, homogeneity of variance, and multicollinearity for the sample, and all assumptions were assessed and met.

Descriptive results show that 18% of first-present patients tested positive for COVID-19 using PCR testing, and 82% of patients tested negative. When comparing between the cohorts testing positive and negative for COVID-19, there are measurable differences between the means for most of the WBC biomarkers (Table 1). Changes in RBC biomarkers were smaller but still noticeable in raw descriptive percentages. The full effect of those differences in both WBC and RBC biomarkers between COVID-19 positive and negative patients is best demonstrated in the forthcoming results from the multivariate inferential analyses. The distribution of age between those testing positive and negative was slightly skewed toward the older patients testing positive. The distribution of sex for the total study cohort was 55% female and 45% male. However, in the COVID-19 positive group, males were a greater proportion, accounting for 52% of infected patients. The effect of males having a greater likelihood of testing positive for COVID-19 was a powerful confounding variable in this study.

**Table 1.**
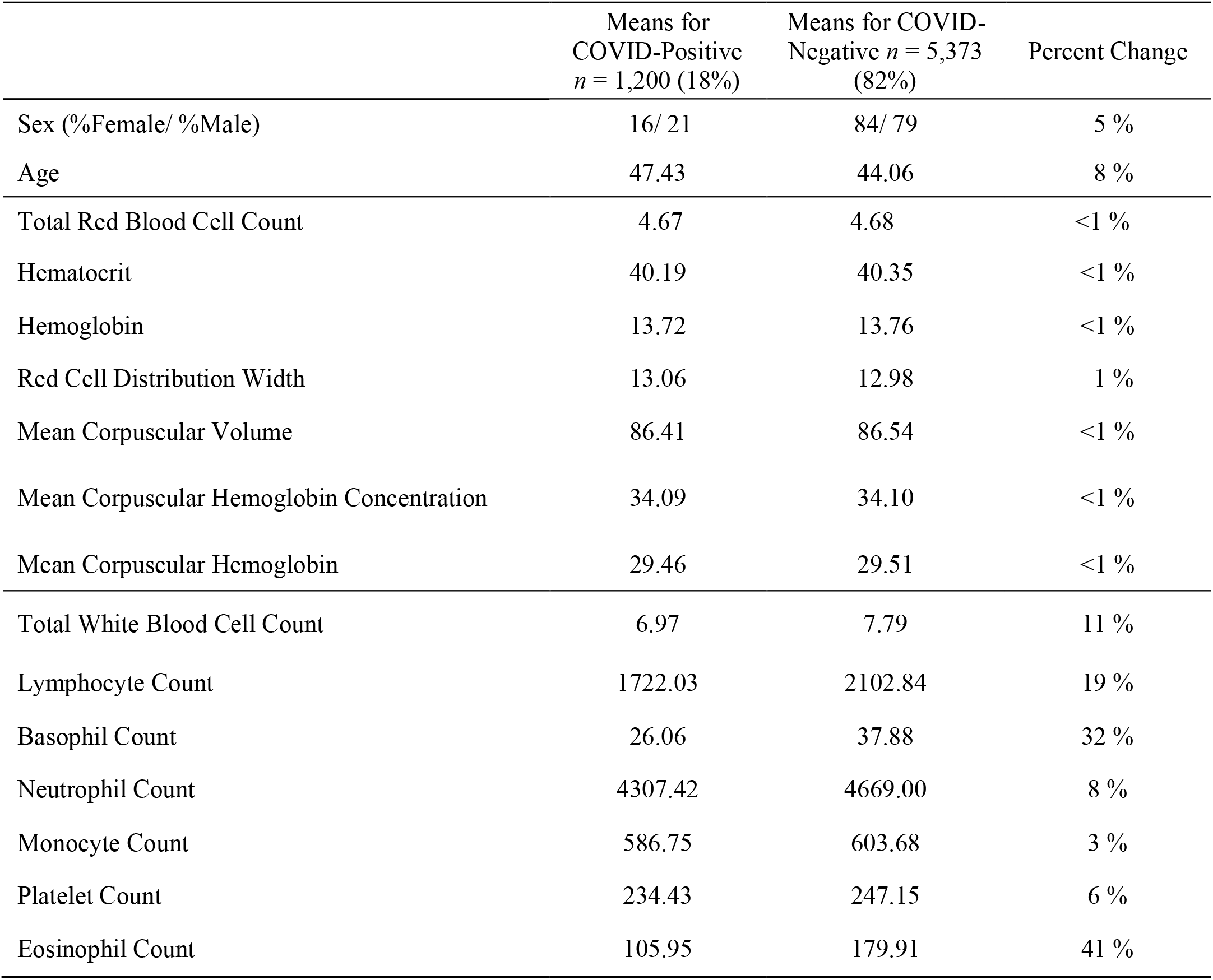
Mean Values and the Percentage Difference for Fourteen Hematological Biomarkers, Age and Sex, for Hospital Patients Testing Positive and Negative for COVID-19 in 2020, Sao Paul, Brazil.

Point biserial correlations were conducted for each of the fourteen hematology biomarkers by the outcome PCR test result (Table 2). At the bivariate level, WBC biomarkers demonstrated statistically significant correlations with PCR test results, but none of the RBC biomarkers were predictive; slight trends were evident but not significant. The demographic variables of age and sex both showed strong statistically significant relationships with PCR test results, with being male appearing to be a significant risk factor for testing positive: OR=1.43 (1.26-1.62). The relationship between patient age and COVID-19 is consistent with the current understanding of the disease impacting the older population in greater proportion. The result of sex of the patient being a powerful predictor is new to the literature.

**Table 2.**
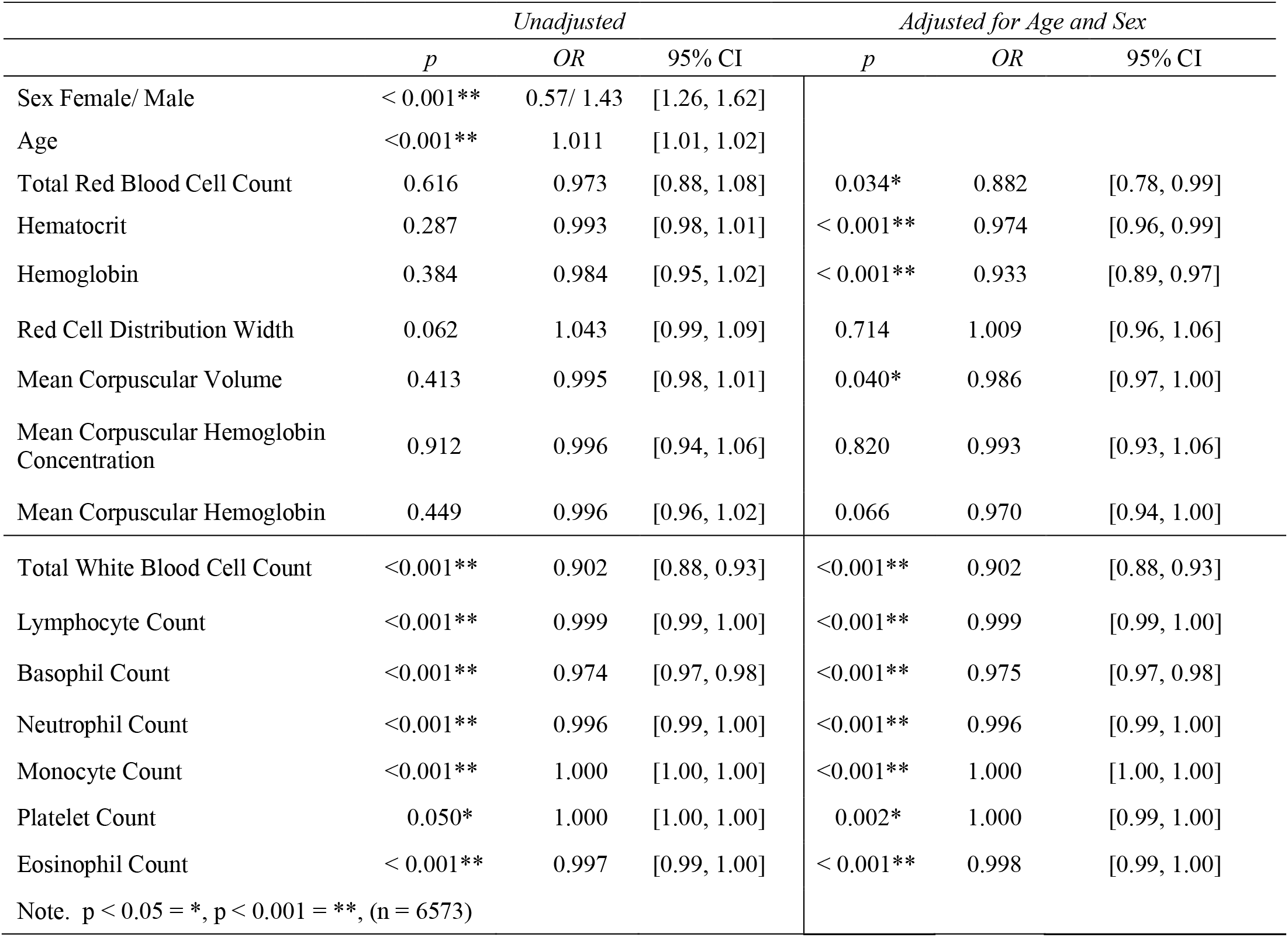
Odds Ratios for Fourteen Hematological Biomarkers Predicting COVID-19 PCR Test Results for First-Present Patients in 2020, Sao Paulo, Brazil: Unadjusted and Adjusted for Age and Sex.

|  | *Unadjusted* |  |  | *Adjusted for Age and Sex* |  |  |
| --- | --- | --- | --- | --- | --- | --- |
|  | *p* | *OR* | *95% CI* | *p* | *OR* | *95% CI* |
| Sex Female/ Male | < 0.001** | 0.57/ 1.43 | [1.26, 1.62] |  |  |  |
| Age | <0.001** | 1.011 | [1.01, 1.02] |  |  |  |
| Total Red Blood Cell Count | 0.616 | 0.973 | [0.88, 1.08] | 0.034* | 0.882 | [0.78, 0.99] |
| Hematocrit | 0.287 | 0.993 | [0.98, 1.01] | < 0.001** | 0.974 | [0.96, 0.99] |
| Hemoglobin | 0.384 | 0.984 | [0.95, 1.02] | < 0.001** | 0.933 | [0.89, 0.97] |
| Red Cell Distribution Width | 0.062 | 1.043 | [0.99, 1.09] | 0.714 | 1.009 | [0.96, 1.06] |
| Mean Corpuscular Volume | 0.413 | 0.995 | [0.98, 1.01] | 0.040* | 0.986 | [0.97, 1.00] |
| Mean Corpuscular Hemoglobin Concentration | 0.912 | 0.996 | [0.94, 1.06] | 0.820 | 0.993 | [0.93, 1.06] |
| Mean Corpuscular Hemoglobin | 0.449 | 0.996 | [0.96, 1.02] | 0.066 | 0.970 | [0.94, 1.00] |
| Total White Blood Cell Count | <0.001** | 0.902 | [0.88, 0.93] | <0.001** | 0.902 | [0.88, 0.93] |
| Lymphocyte Count | <0.001** | 0.999 | [0.99, 1.00] | <0.001** | 0.999 | [0.99, 1.00] |
| Basophil Count | <0.001** | 0.974 | [0.97, 0.98] | <0.001** | 0.975 | [0.97, 0.98] |
| Neutrophil Count | <0.001** | 0.996 | [0.99, 1.00] | <0.001** | 0.996 | [0.99, 1.00] |
| Monocyte Count | <0.001** | 1.000 | [1.00, 1.00] | <0.001** | 1.000 | [1.00, 1.00] |
| Platelet Count | 0.050* | 1.000 | [1.00, 1.00] | 0.002* | 1.000 | [0.99, 1.00] |
| Eosinophil Count | < 0.001** | 0.997 | [0.99, 1.00] | < 0.001** | 0.998 | [0.99, 1.00] |
| Note. | p < 0.05 = *, p < 0.001 = **, (n = 6573) |  |  |  |  |  |

When logistic regressions were conducted for each of the fourteen hematology biomarkers while adjusting for the demographic variables of age and sex, the strength of association with PCR results increased for most of the hematology biomarkers (Table 2). Some biomarkers were weak predictors when analyzed at the bivariate level, but adjusting for age and sex improved the model so that most of the hematology biomarkers demonstrated significant relationships.

Overall results from the adjusted models found that ten of the fourteen hematology biomarkers were statistically significant predictors of the COVID-19 PCR test result. Five RBC and WBC biomarkers demonstrated the strongest relationship; Total White Blood Cells: OR=0.90 (0.88-0.93), Basophil: OR=0.97 (0.97-0.98), Total Red Blood Cell: OR=0.88 (0.78-0.99), Hemoglobin: OR=0.93 (0.89-0.97), Hematocrit: OR=0.97 (0.96-0.99). Only three hematology biomarkers demonstrated no statistically significant association with COVID-19 test results, and all three are RBC indices: Red Cell Distribution Width, Mean Corpuscular Hemoglobin Concentration, and Mean Corpuscular Hemoglobin. Although these RBC indices were not statistically significant, they did show trends that mirror the results expressed in the other biomarkers.

During an infection, similar changes in leukocytic cells are expected across all patients presenting similar symptoms. However, results here show immune responses were differential between the COVID-19 positive and negative patients across all White Blood Cell biomarkers (Table 2). All seven WBC biomarkers demonstrated statistically significant relationships with PCR test results, two of which had strong predictive effects. Two WBC indices (Monocytes and Platelets) were predictive of PCR test results, yet they produced ORs equal to 1.000. The levels of Monocytes and Platelets measured in the blood are statistically significant predictors of the disease state as they are not due to chance alone, but blood levels alone are not predictive of the outcome because the ORs are equal to 1.00.

The results reported in this study were produced with CBC data run at the interval level for each biomarker, as reported by the hospital. A secondary analysis was conducted in which the values for the fourteen CBC biomarkers were converted to nominal variables, with values assigned as either normal or abnormal, as defined by the CBC reference ranges. When CBC data was examined as either within normal or abnormal ranges, the predictive effect of the hematology values was evident, but at much lower levels than when analyses were conducted with CBC data at the interval level of measure. This exercise revealed that much of the predictive power of association occurred within the ranges defined as normal for CBC hematological biomarker levels, and that a review of CBC results with the standard lens of normal/abnormal, would not present the true impacts of the virus on the full blood profile.

## Discussion

Overall results suggest the SARS-CoV-2 virus produces a clear biosignature in both the red and white blood cell lines that is measurable using the Complete Blood Count derived from a routine blood draw. If confirmed, this relationship could offer a faster probabilistic predictor of COVID-19 infections for patients when they first-present at hospitals. Most healthcare facilities have the resources to run a standard Complete Blood Count within an hour via a hematology analyzer; conducting a CBC test is routine practice in most healthcare settings.

Studies are reporting abnormalities for certain hematology biomarkers in their COVID-19 patients, many of whom were in the later phases of disease progression.^7 ,26^ This study is the second to compare CBC hematology results between COVID-19 patients and non-COVID-19 patients, and both studies report statistically significant differential hematological responses in sick patients with similar symptoms.^19^ This current study expanded the profile of biomarkers and demographics considered by examining the RBC biomarkers and the patients’ sex, and only considered data from the early phase of the disease. This investigation of all fourteen hematology biomarkers for both red and white blood cells is a more comprehensive assessment of how the full hematology profile is impacted by COVID-19. The inclusion of patient sex also showed it to be an important variable of consideration due to the unexpected effect between males and females.

Results from the RBC indices are concerning because their primary function is not related to immunological responses. Some studies have reported abnormal RBC results in individual indices in their COVID-19 patients.^31,30,29^ This study examined all seven of the RBC biomarkers while adjusting for age and sex. Results suggest even at the early phase of the disease, erythrocyte functions are measurably impacted.

Three of the five hematology biomarkers that were the strongest predictors of PCR test results were erythrocytic (Table 2). Erythrocytes are responsible for oxygen transport, so even a 1% change in availability can have systemic affects. Results for Total RBC counts show the lower they are, the greater the probability of the patient testing positive. Hematocrit (HCT) is the proportion (by volume) of the blood that consists of red blood cells. Results show the lower the HCT, the greater the probability of the patient testing positive. Hemoglobin (HGB) is the oxygen-transporting protein found in RBCs. HGB contains a heme group for binding oxygen and transporting to the body’s cells. Results show that the lower the HGB, the greater the probability of the patient testing positive. The other three RBC biomarkers demonstrated similar trends, but the associations were not statistically significant. The effect of COVID-19 on erythrocyte function reported in this study suggests the virus impacts the RBCs ability to transport oxygen, even at the early phases of the disease. All living cells require oxygen, and decreased availability can trigger rapid clinical decline of a patient, with a wide range of negative health outcomes including organ failures and death.

White Blood Cells are the human body’s immunological response system, and although only certain WBC biomarkers should be affected during a viral infection, this study reports that even at the early phase of the disease, all leukocyte functions are measurably impacted differently from other sick patients who do not have COVID-19. Two WBC biomarkers expressed the strongest predictive values for PCR test results: Total WBC counts and Basophil counts. As anticipated, lower Total WBC counts in the blood increased the probability of the patient testing positive, as has been reported by others who studied populations of known COVID-19 positive patients.^1,15^ Unexpectedly, lower levels of Basophils (granulocytic) were also significantly associated with the probability of testing positive. Basophils are primarily associated with allergies and parasitic infections, and they comprise less than 1% of leukocytes. Basophils are the only circulating leukocytes that contain histamine, and they are a source of the major Th2-driving cytokine, IL-4, early in immune responses. There are reports that basophils appear to have an active role in the immune response to COVID-19.^21,23^ The other five WBC biomarkers also showed statistically significant predictive values, but with less power. Results for four of those biomarkers are unusual because neutrophils, monocytes, platelets, and eosinophils are not typically activated during a viral infection because they have other primary functions in the immune system.

In this study, only 18% of first-presentation patients tested positive for COVID-19. This percentage is supported by similar reports in other regions where positivity rates range from 8% to 25%.^13^ This result highlights the important need to rapidly identify and isolate the infected patients so the other 80% are not inadvertently exposed.

Age was a significant predictor of PCR test results, with older patients more likely to test positive. The effect of age was an anticipated result that is supported by the literature.^33^ However, the effect of patient age on testing positive was not as strong as anticipated, suggesting the possibility that comorbidities may be a stronger predictor of infection than age; an area in need of further research.

Patient sex was also a significant predictor of PCR test results, with male patients more likely to test positive for COVID-19 than females. The strong predictive effect of being male is a novel finding, and further investigation is encouraged. The driver for this differential may be socially-based behavioral aspects related to job exposure, etc., or there may be different immunological responses or sex hormones in the blood that make men more susceptible to becoming infected.^9^ A powerful standout finding of this study is that reported low Total Red Blood Cell levels in males was a very strong indicator that the patient was COVID-19 positive.

The findings in this study were tested for validity by developing a preliminary model from the results, and running data collected from a different hospital in Sao Paulo, Brazil. The data for 200 patients were run through the crude model and results predicted the known PCR results with 82% sensitivity and 80% specificity. A more refined model is being developed as data and variables are added, with the intention of making the model publicly available soon in an accompanying article.

Important qualifiers to consider include the data was collected early in the pandemic; none of the patients were infected with either the Delta or Omicron variants. Data were collected from one hospital in one country, and results need to be reproduced in other populations to test external validity. The study method is reproduceable and has strong internal validity because all quantified data are collected by third parties following standardized international protocols.

A routine CBC test may be an additional tool for front-line healthcare facilities, but the test alone should not be the only diagnostic tool when PCR is available. PCR testing provides the most valuable data through direct identification of viral RNA, including potential sequencing on type of variate. CBC test results may provide a fast first probabilistic indicator of COVID-19 to be used in conjunction with comprehensive metabolic panels, inflammatory marker analysis, and ultimately PCR testing for confirmation and strain identification. Collectively, the additional testing protocol would provide a larger profile of quantified results to aid fast and accurate diagnosis and isolation.

### Public Health Implications

Results demonstrate a routine CBC hematology test may be an effective diagnostic tool for probabilistic identification of COVID-19 infection. This finding would increase the COVID-19 testing speed and availability for healthcare systems with limited resources. With less than 20% of first-present patients testing positive, fast accurate testing is important for proper isolation to reduce further transmission to the other patients. Findings also indicate the virus measurably impacts a range of hematology biomarkers including vital red blood cell attributes. These findings demonstrate the virus has a wider range of hematology impact than reported to date. Results from this study are being used to develop a predictive model based on CBC test results with the level of sensitivity and specificity equal to PCR technology. Open access to the model will improve patient triage and isolation.

## Data Availability

All data produced in the present study are available upon reasonable request to the authors

## Attachments

1. Araya S, Wordofa M, Mamo MA, et al. The magnitude of hematological abnormalities among covid-19 patients in Addis Ababa, Ethiopia. J Multidiscip Healthc. 2021;14:545–554. doi:10.2147/JMDH.S295432

2. Shen L, Cui S, Zhang D, Lin C, Chen L, Wang Q. Comparison of four commercial RT-PCR diagnostic kits for COVID-19 in China. J Clin Lab Anal. 2021;35(1):1–8. doi:10.1002/jcla.23605

3. Magno L, Rossi TA, de Mendonça-Lima FW, et al. Challenges and proposals for scaling up COVID-19 testing and diagnosis in Brazil. Cienc e Saude Coletiva. 2020;25(9):3355–3364. doi:10.1590/1413-81232020259.17812020

4. Ramanathan K, Antognini D, Combes A, et al. Clinical impact of molecular point-of-care testing for suspected COVID-19 in hospital (COV-19POC): a prospective, interventional, non-randomised, controlled study. 2020;(January):19–21.

5. Roy Choudhury SH, Shahi PK, Sharma S, Dhar R. Utility of chest radiography on admission for initial triaging of COVID-19 in symptomatic patients. ERJ Open Res. 2020;6(3):00357–02020. doi:10.1183/23120541.00357-2020

6. Jöbges S, Vinay R, Luyckx VA, Biller-Andorno N. Recommendations on COVID-19 triage: international comparison and ethical analysis. Bioethics. 2020;34(9):948–959. doi:10.1111/bioe.12805

7. Waris A, Din M, Khalid A, et al. Evaluation of hematological parameters as an indicator of disease severity in Covid-19 patients: Pakistan’s experience. J Clin Lab Anal. 2021;35(6):1–10. doi:10.1002/jcla.23809

8. Barrett J, Painter H, Rajgopal A, et al. Increase in disseminated TB during the COVID-19 pandemic Taking action to improve post-TB lung health. Int J Tuberc Lung Dis. 2021;25(2):160–161.

9. Bwire GM. Coronavirus: Why Men are More Vulnerable to Covid-19 Than Women? SN Compr Clin Med. 2020;2(7):874–876. doi:10.1007/s42399-020-00341-w

10. Mello LE, Suman A, Medeiros CB, et al. Opening Brazilian COVID-19 patient data to support world research on pandemics. Zenodo. Published online 2020. https://zenodo.org/record/3966427

11. Alves JD, Abade AS, Peres WP, Borges JE, Santos SM, Scholze AR. Impact of COVID-19 on the indigenous population of Brazil: A geo-epidemiological study. Epidemiol Infect. Published online 2021. doi:10.1017/S0950268821001849

12. Silva DI, Silva SB, Magalhaes VCR. Impacto Do Covid‐19 E a Coinfecção Tb/Hiv Em Um Centro De Referência Do Sudeste Brasileiro. Brazilian J Infect Dis. 2021;25(S 1):101277. doi:10.1016/j.bjid.2020.101277

13. Hannah Ritchie, Edouard Mathieu, Lucas Rodés-Guirao, Cameron Appel, Charlie Giattino, Esteban Ortiz-Ospina, Joe Hasell, Bobbie Macdonald DB and MR. Coronavirus Pandemic (COVID-19). Our World Data. Published online 2020. https://ourworldindata.org/coronavirus

14. Valenstein P. Laboratory turnaround time. Am J Clin Pathol. 1996;105(6):676–688. doi:10.1093/ajcp/105.6.676

15. Zhao L, Xing H, Xu L. Effect of SARS-associated coronavirus on peripheral blood picture and liver function. Chinese Crit care Med. 2004;16(11):660–663. http://europepmc.org/abstract/MED/15535899

16. Wang K, Chen W, Zhang Z, et al. CD147-spike protein is a novel route for SARS-CoV-2 infection to host cells. Signal Transduct Target Ther. 2020;5(1):1–11. doi:10.1038/s41392-020-00426-x

17. Stamm P, Sagoschen I, Weise K, et al. Influenza and RSV incidence during COVID-19 pandemic—an observational study from in-hospital point-of-care testing. Med Microbiol Immunol. 2021;210(5-6):277–282. doi:10.1007/s00430-021-00720-7

18. Fenizia C, Galbiati S, Vanetti C, et al. SARS-CoV-2 entry: At the crossroads of CD147 and ACE2. Cells. 2021;10(6). doi:10.3390/cells10061434

19. AlJame M, Imtiaz A, Ahmad I, Mohammed A. Deep forest model for diagnosing COVID-19 from routine blood tests. Sci Rep. 2021;11(1):1–12. doi:10.1038/s41598-021-95957-w

20. Asaduzzaman M, Hossain N, Abul M. Immune response in COVID-19: A review. 2020;(January).

21. Palladino M. Complete blood count alterations in covid-19 patients: A narrative review. Biochem Medica. 2021;31(3):1–13. doi:10.11613/BM.2021.030501

22. Mousavi SA, Rad S, Rostami T, et al. Hematologic predictors of mortality in hospitalized patients with COVID-19: a comparative study. Hematol (United Kingdom). 2020;25(1):383–388. doi:10.1080/16078454.2020.1833435

23. Murdaca G, Di Gioacchino M, Greco M, et al. Basophils and mast cells in COVID-19 pathogenesis. Cells. 2021;10(10):1–13. doi:10.3390/cells10102754

24. Formica V, Minieri M, Bernardini S, et al. Complete blood count might help to identify subjects with high probability of testing positive to SARS-CoV-2. Clin Med J R Coll Physicians London. 2020;20(4). doi:10.7861/CLINMED.2020-0373

25. Arun Prabhakaran Nair, Ashraf Soliman, Muna A. Al Masalamani VDS. View of Clinical Outcome of Eosinophilia in Patients with COVID-19_ A Controlled Study.pdf. Published online 2020:Vol. 91. doi:10.23750/abm.v91i4.10564

26. Karimi F, Vaezi AA, Qorbani M, et al. Clinical and laboratory findings in COVID-19 adult hospitalized patients from Alborz province / Iran: comparison of rRT-PCR positive and negative. BMC Infect Dis. 2021;21(1):1–9. doi:10.1186/s12879-021-05948-5

27. Erdinc B, Sahni S, Gotlieb V. Hematological manifestations and complications of COVID-19. Adv Clin Exp Med. 2021;30(1):101–107. doi:10.17219/ACEM/130604

28. Koupenova M. Potential role of platelets in COVID‐19: Implications for thrombosis. Res Pract Thromb Haemost. 2020;4(5):737–740. doi:10.1002/rth2.12397

29. Berzuini A, Bianco C, Migliorini AC, Maggioni M, Valenti L, Prati D. Red blood cell morphology in patients with COVID-19-related anaemia. Blood Transfus. 2021;19(1):34–36. doi:10.2450/2020.0242-20

30. Diederich L, Iv TCSK, Kuhn V, Kramer CM. Red Blood Cell Function and Dysfunction: 2017;26(13):718–742. doi:10.1089/ars.2016.6954

31. Maheshwari A, Priya G, Bajpai M. Isolated erythrocytosis: A consequence of COVID-19 induced hypoxia. Int J Lab Hematol. 2021;(August):1–2. doi:10.1111/ijlh.13699

32. Ehrenreich H, Weissenborn K, Begemann M, et al. Erythropoietin as candidate for supportive treatment of severe COVID-19. Mol Med. 2020;26(1). doi:10.1186/s10020-020-00186-y

33. Amber L. Mueller, Maeve S. McNamara, David A. Sinclair. Why does COVID-19 disproportionately affect older people? Aging (Albany NY). 2020;12(10):9959–9981.

34. Nikolopoulou GB, Maltezou HC. COVID-19 in children : where do we stand? Arch Med Res. 2021;(July).

35. Sanyaolu A, Okorie C, Marinkovic A, et al. Comorbidity and its Impact on Patients with COVID-19. SN Compr Clin Med. 2020;2(8):1069–1076. doi:10.1007/s42399-020-00363-4

